# Evaluating the Risk of Psilocybin for the Treatment of Bipolar Depression: A Review of the Research Literature and Published Case Studies

**DOI:** 10.1101/2021.04.02.21254838

**Authors:** David E. Gard, Mollie M. Pleet, Ellen R. Bradley, Andrew Penn, Matthew L. Gallenstein, Lauren S. Riley, Meghan DellaCrosse, Emily Garfinkle, Erin E. Michalak, Joshua D. Woolley

## Abstract

Growing evidence suggests that psilocybin, the active ingredient in hallucinogenic mushrooms, can rapidly and durably improve symptoms of depression, leading to recent breakthrough status designation by the FDA and legalization for mental health treatment in some jurisdictions. Depression in bipolar disorder is associated with significant morbidity and has few effective treatments. However, there is little available scientific data on the risk of psilocybin use in people with bipolar disorder. Individuals with bipolar disorder have been excluded from modern clinical trials, out of understandable concerns of activating mania or worsening the illness course. As psilocybin becomes more available, people with these disorders will likely seek psilocybin treatment for depression and have likely already been doing so in unregulated settings. Our goal here is to summarize the known risks of psilocybin use (and similar substances) in bipolar disorder and to systematically evaluate examples of published case history data, in order to critically evaluate the relative risk of psilocybin as a treatment for bipolar depression. We found 17 cases suggesting that there is potential risk for activating a manic episode, thereby warranting caution. Nonetheless, the relative lack of systematic data or common case examples indicating risk appears to show that a cautious trial, using modern trial methods focusing on appropriate ‘set’ and ‘setting’, targeted at those lowest at risk for mania in the bipolar spectrum (e.g., bipolar 2 disorder), is very much needed, especially given the degree to which depression impacts this population.

## INTRODUCTION

Individuals with bipolar disorder experience high suicide rates, decreased quality of life, and impaired overall functioning relative to people without bipolar disorder, and in many cases, individuals with other psychiatric conditions (Dome et al., 2019; Michalak et al., 2008). Further, most negative outcomes are related to the depressive phase of the illness (Calabrese et al., 2004). Unfortunately, current pharmacological treatment options for depression in bipolar disorder are limited (Yalin and Young, 2020). One promising new treatment for unipolar depression is psilocybin, a psychedelic compound that has recently received the designation of ‘Breakthrough Therapy for depression’ by the Food and Drug Administration (Reiff et al., 2020). Given the effectiveness of psilocybin, and the limited treatment options for depressive symptoms in bipolar disorder, we and others are looking to develop a treatment trial for depression in this population. However, little is known about the safety of psilocybin therapy for people with bipolar disorder. Thus, the present study investigated the known risks of psilocybin use in people with bipolar disorder.

Psilocybin is a hallucinogenic compound found in several species of mushrooms that has been used in religious and healing practices by Indigenous peoples for millennia (Hofmann, 2009). While there has been some research evidence in humans for its benefit since the 1950’s, in 1970, the Controlled Substances Act essentially ended psychedelic research in humans (Reiff et al., 2020). Following this period, general societal views of psychedelic use were quite critical, focusing on recreational use, sometimes bordering on hyperbole and sensationalism (Belouin and Henningfield, 2018). This is in contrast to self-report (Carhart-Harris and Nutt, 2010; van Amsterdam et al., 2011), observational (Bouso et al., 2012), and epidemiological data (Hendricks et al., 2015) in contemporary research, which suggests that use of these drugs presents a lower risk of harm than other commonly available substances, and may be associated with mental health benefits. In fact, over the past fifteen years, research has indicated that a single dose of psilocybin in a treatment context may dramatically improve symptoms in major depressive disorder and treatment resistant depression (Carhart-Harris et al., 2016; Galvão-Coelho et al., 2021; Ross et al., 2016), obsessive-compulsive disorder (Moreno et al., 2006), anxiety (Grob et al., 2011), and demoralization in long-term AIDS survivors (Anderson et al., 2020). Though acute psychedelic effects of the drug resolve within four to five hours of oral administration, psilocybin’s beneficial effects on mood and well-being may persist for weeks to possibly months (Griffiths et al., 2016).

Importantly, modern psilocybin trials reflect an appreciation for optimizing the conditions under which the drug is administered, both to reduce risk of adverse events and to increase the likelihood that participants will experience lasting positive effects (Johnson et al., 2008). Specifically, modern trials have emphasized ‘set’ and ‘setting’, which are the procedures that explicitly address the mindset of participants before psilocybin exposure, and the environmental setting in which dosing and post-dosing psychological processing of the psilocybin experience occurs. Sufficient preparation for the psychedelic experience, including rapport-building with study staff, attention to the physical space to enhance comfort and safety, and post-drug session meetings to discuss the experience, are now considered essential best practices (Bogenschutz and Ross, 2016; Johnson et al., 2008). Perhaps because of these conditions and careful screening and structure, modern psilocybin trials have not reported any serious adverse events (Andersen et al., 2021).

### What is known about risks in bipolar disorder and psilocybin and psychedelics?

All modern clinical trials with psychedelics have excluded individuals with bipolar disorder and most have excluded individuals with a family history of bipolar disorder out of a concern for precipitating a manic episode or worsening the course of the condition (Studerus et al., 2011). The scientific evidence for these exclusions, however, is rarely clarified. Nonetheless, there are several rationales for excluding these individuals including: the powerful serotonergic activation from psychedelic substances possibly inducing a Treatment Emergent Affective Switch (TEAS; i.e., the activation of a manic episode through the use of an antidepressant), the potential for adverse events in a population that is at higher risk for impulsive behaviors, as well as clinical consensus drawn from examples in psychiatric and emergency admissions and case reports.

### Psilocybin and TEAS

While the phenomenon of mood polarity switching with antidepressants has been extensively studied and correlations are well documented (Melhuish Beaupre et al., 2020), the biologic mechanism of antidepressant-induced TEAS (and for that matter, mood switching in general) remains poorly understood (Salvadore et al., 2010). Substantial evidence suggests that any effective antidepressant, including glucocorticoids, dopamine agonists, ketamine, and transcranial magnetic stimulation, can induce TEASs (Rachid, 2017; Salvadore et al., 2010; Tondo et al., 2010; Yamaguchi et al., 2018). However, the risk for TEASs appears to be higher in response to serotonergic antidepressants, such as SSRIs, SNRIs, and most tricyclic antidepressants (TCAs) (Leverich et al., 2006; Peet, 1994; Post et al., 2006). Consequently, the use of such serotonergic agents in the treatment of bipolar depression is controversial, leading some to recommend avoiding their use whenever possible (Ghaemi et al., 2003). On the other hand, others argue that the risk of TEAS may be overstated (Möller and Grunze, 2000) at least partly because one study found that TEAS are more common in patients with bipolar treated with TCAs (11.2% with a TEAS) compared to SSRIs (3.7% with a TEAS), which are more commonly used now (Peet, 1994). Further complicating matters is evidence that serotonergic antidepressants in patients with bipolar 2 may be effective for treating depression without inducing enduring switches of mood polarity (Altshuler et al., 2017; Amsterdam and Shults, 2005), but may cause an increase in short term changes in mood reflecting mixed states or rapid cycling (Amsterdam and Shults, 2010). In sum, little is understood about the mechanisms of TEASs and there is active debate about the risk and benefits of using serotonergic agents in bipolar disorder with outstanding issues including characterization of the disorder (e.g., type 1 vs 2), the type of agent (e.g., TCA vs. SSRI), and the concomitant use of mood stabilizers. Given these uncertainties, it is challenging to predict whether a psychedelic such as psilocybin is likely to induce a TEAS.

Though the risk of TEAS after a single psychedelic dose is currently unknown, there are reasons to be concerned. First, psilocybin is a potent antidepressant and all antidepressants carry some risk of inducing TEAS (Rachid, 2017; Salvadore et al., 2010; Tondo et al., 2010; Yamaguchi et al., 2018). Second, psilocin, the active metabolite of psilocybin, is a serotonin transporter inhibitor and 5-HT2A receptor partial agonist that also binds to the 5-HT2C, 5-HT1A, and 5-HT1B receptors (Blough et al., 2014; Johnson et al., 2019; Rickli et al., 2016). While different from standard antidepressants in many ways, psilocin has serotonergic mechanisms of action, which could theoretically promote TEASs (Leverich et al., 2006; Peet, 1994; Post et al., 2006). Third, the relationship between antidepressant drug administration regimen and TEAS is unknown. Psilocybin is typically administered in a distinctly different way from standard antidepressants. Standard antidepressants are dosed daily, with generally imperceptible acute effects on mood. They appear to act gradually, often taking weeks or more to demonstrate clinically significant benefit. Psilocybin treatments, in contrast, usually involve a single high-dose administration of the drug that produces profound subjective effects during the period of intoxication. Antidepressant effects are apparent within hours after dosing, and preliminary evidence suggests that these improvements can persist for weeks to months without further administration of the drug. Whether this dramatically different approach to dosing--and its impact on serotonin signaling--increases or decreases the risk of TEAS relative to standard antidepressants has yet to be determined.

### Adverse events and research trials

While all individuals with bipolar disorder and most individuals with a first degree relative with bipolar disorder have been excluded from recent psychedelic research, given the large and growing literature in this area, it is certainly possible that individuals who did not know that they or a family member had bipolar disorder, or who had yet to meet the criteria for the disorder, have been included in previous studies. However, with the structures of ‘set’ and ‘setting’ in place, and a careful attention to participant well-being, participants have not experienced serious adverse events in these studies. For example, in an analysis of 110 healthy participants who completed a total of 227 laboratory-based psilocybin administration sessions, there were no instances of prolonged psychosis, mania, persisting perceptual changes, or other long-term functional impairment in any participants (Studerus et al., 2011). In a systematic review of psychedelics in modern and pre-prohibition psychedelic studies of psilocybin, lysergic acid diethylamide (LSD), and ayahuasca, no cases of severe adverse events (defined as prolonged psychosis, or Hallucinogen Persisting Perception Disorder; HPPD) were found (Rucker et al., 2018). Early research on LSD and other psychedelics did in fact attempt to treat individuals who likely had bipolar disorder. For example, Busch and Johnson (1950) described three individuals with “manic depression” who were treated with LSD. While there was no reported follow up on how the patients fared, indications were that while under the influence of LSD, all three patients were more active and agitated, although it was not clear if this was compared to other patients. One 57-year-old woman with “chronic mania” spoke more rapidly and emotionally after a “small” LSD dose, a second 48-year-old woman with “manic depression” was given LSD during a manic phase and was described to be louder and combative, and a third 49-year-old woman with mania was described as more talkative, irritable, and suspicious. Unfortunately, the lack of follow-up (and other methodological problems with pre-prohibition studies) makes it impossible to disentangle the immediate effects of the substance from what might have occurred later, including any worsening of the underlying illness (Rucker et al., 2018).

### Epidemiology research on psychedelic use

The analysis of recreational hallucinogen use in the general population may provide additional insight into the risk of psilocybin and other psychedelics in people with bipolar disorder, given that the large sample sizes likely include individuals with bipolar disorder or a family history of bipolar disorder. For example, a large-scale survey of more than 130,000 participants indicated that hallucinogen use (in approximately 22,000 individuals) was not a predictor for subsequent mania, psychosis, or mental health treatment (Johansen and Krebs, 2015). In another large-scale survey of over 190,000 participants, individuals who had used psychedelics were at *reduced* odds of psychological distress and suicidality (while participants who reported other non-psychedelic drug use had increased risk of these factors) (Hendricks et al., 2015). While these studies focused on drug use broadly, one study specifically focused on community participants who had ‘challenging or difficult experiences’ while on psilocybin and other hallucinogens. Of 1,900 respondents, 2.6% reported behaving in a physically aggressive or violent manner while on the substance, and three respondents reported enduring psychotic symptoms (Carbonaro et al., 2016); these accounts could potentially reflect activation of a manic episode or the onset of a bipolar illness. However, there were no clear reports of enduring symptoms of mania or activated bipolar illness. Finally, researchers looked at the number of cases of psychotic and bipolar disorders subsequent to over 130,000 doses of ayahuasca given between 1994-2007 at the União do Vegetal (UDV) church in Brazil (Lima and Tofoli, 2011). In total there were 29 cases of psychotic diagnoses, of which 4 were labeled ‘bipolar affective disorder; psychotic manic episode.’ These researchers concluded that the number of psychosis and other cases from this sample is slightly less than expected in the base rate of the population. This is somewhat surprising given that many ayahuasca retreats involve other aspects that might increase the risk of a manic episode (including decreased sleep and repeated dosing). Overall these epidemiological studies do not appear to indicate a clear risk of mania with psychedelic use. Considering that these epidemiological studies assess risk by calculating the overall number of adverse cases divided by the total number of uses, they shed some light on what the denominator may be in the risk evaluation. That is, even in large samples of psychedelic use, there are only a handful of adverse events. In addition, large scale surveys of drug use indicate that between 8-10% of the population (or over 26 million people in the US) reports using psilocybin (or other psychedelics) in their lifetime (Center for Behavioral Health Statistics and Quality, 2016). Thus, while the numerator of the equation is not clearly known, the denominator is quite large.

#### Review of Published Cases

Given the relative dearth of information on adverse events in bipolar disorder within the broader psychedelic literature, we turned our attention to the case study literature. Our goal here was to critically explore these cases for any adverse effects of psilocybin (and related psychedelics) in individuals with bipolar disorder, and the potential for these substances to activate a manic episode. We focused this portion of the review on psilocybin as the current evidence suggests that this compound may be effective in treating depression. However, there is a limited number of cases indicating specific risk associated with psilocybin use in bipolar disorder. Thus, we broadened our question of risk to include all serotonergic psychedelic tryptamines including LSD, ayahuasca, and DMT because these compounds share similar molecular (and experiential) characteristics (dos Santos and Hallak, 2020; Nichols, 2016). Also, because it can be difficult to clinically differentiate a manic from a psychotic episode, we included case histories and descriptions of what appear to be manic *or* psychotic behavior with manic behavior that persisted beyond the immediate effects of the substance. We both broadened the search to include all serotonergic psychedelics and to manic-like behavior to take the most cautious approach, in order to find as many cases as possible. Importantly, the focus of this review was on the risk of activating manic episodes in individuals with bipolar spectrum disorder, and not on the risk of activating a psychotic episode leading to a diagnosis of a psychotic spectrum disorder. Therefore, we focus here on the former.

## METHOD

### Identification and selection of publications

A comprehensive search of case studies published through June 2021 was conducted using the following electronic databases: PubMed, Web of Science, and PsychInfo. Our initial search focused specifically on psilocybin and included the following terms ((“psilocybin”) OR (“hallucinogenic mushrooms”) OR (“magic mushrooms”)) AND ((“case study”) OR (“case report”) OR (“case review”)). This search revealed a small number of publications (Pubmed=68, Web of Science=17, PsychInfo=8) prior to inclusion/exclusion criteria below (which narrowed the case studies down to (Pubmed=5, Web of Science=3, PsychInfo=2, or a total of five non-overlapping cases), and so we expanded the search to include all classic psychedelics (((“psilocybin”) OR (“hallucinogenic mushrooms”) OR (“magic mushrooms”) OR (“LSD”) OR (“ayahuasca”) OR (“DMT”)) AND ((“case study”) OR (“case report”) OR (“case review”)).

### Additional inclusion/exclusion criteria

Case studies were included if they met the following criteria: 1) written or translated into English with at least a DOI or abstract to locate the article, 2) described an individual who had clearly taken a psychedelic, 3) included individuals with persistent effects from the substance that was adverse: 3A) For individuals who had a diagnosis of bipolar disorder, or in examples where the authors described a history that indicated a likely diagnosis of bipolar disorder, we included these individuals if there were *any* adverse events after taking a psychedelic substance, 3B) for individuals who did not have a clear diagnosis or history of bipolar disorder (i.e., the vast majority of cases), we included the case if the problems or symptoms related to the adverse event were described in a way that could be interpreted as mania (e.g., increased goal-directed behavior, decreased need for sleep, etc.) or positive symptoms of psychosis (e.g., hallucinations or delusions) that continued beyond immediate, intoxication effects of the substance, 4) following DSM-5 criteria (*Diagnostic and Statistical Manual of Mental Disorders: DSM-5*, 2017) for duration of a manic or hypomanic episode, we included only cases with effects that lasted at least 4 days, or involved hospitalization for the adverse effects, or involved effects that were clearly severe in nature (e.g., resulting in extreme symptoms or problems). 5) We did not exclude polysubstance use, but have noted polysubstance use in the included table (Table 2), 6) while we included psychosis symptoms or adverse events in the initial screening, we excluded cases that resulted in symptoms or problems that did not also ultimately result in mania or hypomania or a diagnosis of a bipolar disorder (e.g., individuals who were subsequently diagnosed with schizophrenia or HPPD).

### Process

Three authors (MP, LR, and MG) independently reviewed all electronic databases. Four authors (LR, MG, DEG, & JW) conferred to make final decisions when it was unclear whether a case met specific criteria (usually duration of the episode (#4) or whether the ultimate diagnosis would be mania or purely psychosis (#6)). As mentioned above, the initial search was expanded to include other classic psychedelics which resulted in a more substantive case number (Pubmed=269, Web of Science=141, PsycInfo=152). After duplicates were removed and screened using the aforementioned criteria, the references in the remaining cases were examined for additional relevant case studies, but no additional cases were found. In total, 44 cases were initially included based on criteria 1-3 (in English or translated, and clear adverse events with symptoms), and then 27 were excluded based on criteria 4-6 (duration of mania was not met, or the result was clearly psychosis or HPPD only), leaving 17 cases. See Figure 1 for a flow chart of inclusion/exclusion.

**Figure 1:**
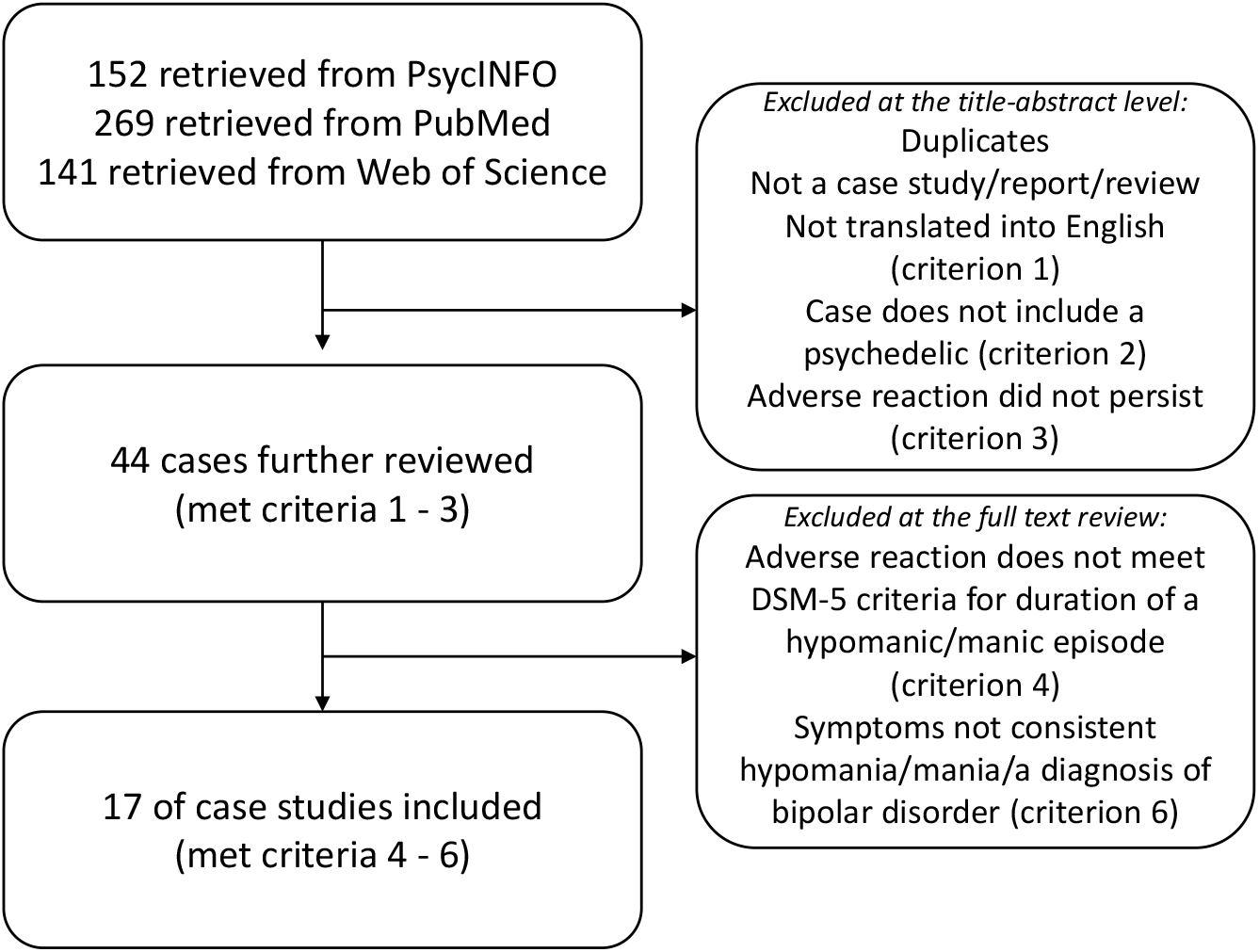
Flow chart for the search and selection of the final cases.

## RESULTS

In total, 17 published case studies met all of the selection criteria, five of which involved psilocybin (Barbic et al., 2020; Bickel et al., 2005; Brown et al., 2017; Cohen, 1966; Davies, 1979; Dewhurst, 1980; Dos Santos et al., 2017; Hyde et al., 1978; Lake et al., 1981; Paterson et al., 2015; Perera et al., 1995; Prajapati et al., 2016; Reich and Hepps, 1972; Sami et al., 2015; Szmulewicz et al., 2015; Hendin & Penn, 2021). Table 1 summarizes the case content (each case is numbered) and Table 2 summarizes the themes in each case. In terms of themes, of the 17, six case studies appeared to involve some history of mania or hypomania before ingestion of the psychedelic, or a family history of bipolar disorder (Cases 7, 9, 11, 13, 15, 16). Five reported some form of additional substance use in the current adverse event (Cases 6, 9, 10, 12, 13), seven had some substance abuse or polysubstance use history prior to the episode (Cases 2, 5, 8, 10, 12, 14, 15), and nine used the substance repeatedly in a relatively short time period (Cases 1, 3, 4, 5, 9, 10, 12, 13, 17). The length of the adverse events varied (often confounded by various treatments) but ranged from a few days to a few months. Most of the case studies noted whether the symptoms had resolved, which was typically within days to weeks, although this, too, was confounded by prescribed treatments. From these cases a number of helpful insights can be gleaned.

**Table 1:** The final 17 cases along with brief descriptions of the cases.

| Case number | Author, year | Brief summary of case |
| --- | --- | --- |
| 1 | Cohen, 1966 | (4th case of 7 'psychotic reactions' in a list of LSD adverse reactions). Age not listed of a man who took LSD 6 or more times and began to believe he was the Savior. He enlisted his wife and other disciples to live in the mountains. At hospitalization he was grandiose and 'hypomaniacal'. Timeline and outcome unclear |
| 2 | Reich & Hepps, 1972 | 22 yo M, using amphetamines, cannabis and LSD over a 2 year period. Had a 'bad trip' on LSD, ultimately attacking an elderly woman. A few months later took LSD again became paranoid and convinced of a Nazi plot, flew home to Israel, withdrew of his money from his bank, flew back to Europe, traveled around and ultimately stabbed a stranger believing him to be a Nazi soldier. |
| 3 | Hyde et al., 1978 | 20 yo M, consumed fresh psilocybin mushrooms multiple times over course of a week, while depriving himself of food and sleep. Took final dose of 50-60 mushrooms, then admitted to hospital within 24 hours of the large dose. Restricted speech, motor stereotypies of saluting. In another 24 hours, became fearful and aggressive, and at 72 hours showed waxy flexibility, fear and agitation. Over next few days, recovered and was cheerful and pleasant, reporting no more psychiatric symptoms. |
| 4 | Davies, 1979 | 26-year old M, began staying up all night to read the Bible, had a number of grandiose delusions, reported that he took consumed dried Psilocybe semilanceata, a couple months prior but not clearly linked. Hospitalized and discharged after 5 weeks, after which he had a few bouts of depression but no more psychotic symptoms. |
| 5 | Dewhurst, 1980 | 25 yo M, took ~200 fresh psilocybin mushrooms, became paranoid, attacked three detectives that arrested him. Hospitalized then re-admitted next day after antidepressant overdose, including aggressive behavior, smashing several windows and attacking the nursing staff. Was hospitalized a third time for 10 weeks, and was discharged and, symptoms remitted and he 'remained well' |
| 6 | Lake et al., 1981 | 31 yo M with some alcohol use problems took LSD one time and had a 'bad trip' with mostly experiences of anxiety and guilt. Two weeks post LSD use his sleep decreased and he became very religious, manic and delusional (no other symptoms of psychosis), ultimately being hospitalized and treated effectively with lithium. |
| 7 | Perera, Ferraro, & Pinto, 1995 | 21 yo F admitted to hospital with elation, decreased need for sleep, paranoia, mood swings, catatonia, echolalia. Symptoms began 1 week prior, after taking LSD at music festival. Mother has bipolar disorder, uncle schizophrenia. treated with lithium and chlorpromazine and was symptom free 3 months later |
| 8 | Bickel et al., 2005 | 25 yo M, presented to ER after ingestion of psychoactive fungi. Was agitated, hallucinating, restless and aggressive. History of heroin, opiate, and cannabis abuse (but reported only using psilocybin mushrooms). After seventh day at hospital, lost sight in both eyes, became agitated, very restless & disoriented. 6 months later client presented to clinic fully orientated w/ no neurological deficits and w/ normal visual abilities and no other symptoms. |
| 9 | Paterson, Darby, & Sandhu, 2015 | 42 yo M, family history of bipolar disorder. 3 weeks before hospitalization, started smoking DMT. Upon admission to hospital, was delusional, thought content was loose, disorganized, paranoid, with grandiose delusions for 12 days. Patient was hyperverbal & intrusive. Was given an antipsychotic and symptoms remitted after 14 days, and continued symptom free at 6 month follow up. |
| 10 | Sami et al., 2015 | 34 yo M history of TBI, heavy use of psychedelics and other substances over 6 month period, Admitted to hospital with psychotic symptoms including grandiose delusions of being Jesus, irritability and flight of ideas. Relapsed 6 months after hospitalization with similar substances |
| 11 | Szmulewicz, Valerio, & Smith, 2015 | 30 yo M arrived 2 days after a 4 day ayahuasca retreat. He had mystical and paranoid and delusional ideas at admission, along with racing thoughts, elevated energy and euphoria. Reported possible hypomanic symptoms two weeks prior. Treated at the hospital with antipsychotics and no symptoms remained one month later. |
| 12 | Prajapati et al., 2016 | 19 yo M brought to ER by mother with visual hallucinations, confusion and hyperarousal. Patient later admitted to LSD use two days earlier, as well as other drug use beginning 5 months prior. Patient remained agitated, confused and at times catatonic. Treated successfully over several days in the hospital with antipsychotic medication |
| 13 | Brown et al., 2017 | 40 yo M, retired psychiatrist w/ Bipolar I. On arrival to ER, patient was nonverbal, combative, & required 6 security guards to restrain him. Symptoms of psychosis and mania. Pressured speech, hyperreligious, and delusional. Patient had been self-medicating to treat refractory depression with DMT and phenelzine. Only had one prior manic episode, and had otherwise remained depressed for the majority of his life. Patient had been taking up to 1g of vaporized DMT daily for past 6 months. Patient still remained hypomanic and discharged home against medical advice. |
| 14 | dos Santos, Bouso, and Hallak, 2017 | 40 yo F attended a 2 day ayahuasca retreat for self improvement. On the second day began to have psychotic symptoms including paranoid ideas, delusions, hyperarousal, and excessive talking. She then had several nights without sleep, and other delusional thoughts at time of hospitalization. |
| 15 | Barbic et al., 2020 | (Case 1 of 2): 25 yo M brought by police to ED due to acute psychosis at DMT use: tangential, pressured speech, delusions of being a fictional character from a movie. Combative with police and paramedics. Family indicated he had been isolating for the past 5 months. Only family psychiatric history was first cousin with bipolar disorder. Psychosis slowly resolved & discharged after 20 days. |
| 16 | Hendin and Penn, 2021 | 21 yo F presented to outpatient psychiatry one month following hospitalization related to episode of mania triggered by psilocybin use. This was her first psilocybin experience, and despite a family h/o bipolar disorder (father, paternal grandmother), patient had no prior manic episode. She reported that psilocybin experience was pleasant and "mystical" but that 36hrs post-dose, she experienced irritability, pressured speech, decreased need for sleep, paranoia, and delusions that led her family to contact the police. She was admitted to inpatient behavioral health and stabilized on lithium and aripiprazole, and later lamotrigine. She later had one irritable mixed episode of mania that has since resolved. |
| 17 | Palma-Alvarez et al., 2021 | 41 yo M with h/o ayahuasca use since age 25 presented to ED experiencing restlessness, unspecified fears, delusions of reference, severe mood fluctuations, sensory-perceptive distortions, somesthetic hallucinations, and simple auditory hallucinations. He had most recently been using ayahuasca alone at home 1x/wk for a month to manage emotional distress following a relationship. Following antipsychotic administration in ED, acute symptoms remitted and patient was discharged to SUD treatment facility. |

**Table 2:** Themes from each of the 17 final cases.

| <u>Case number</u> | <u>Author, year</u> | <u>Length of adverse event</u> | <u>psychedelic substance</u> | <u>polysubstance use during event/prior</u> | <u>Mania &/or psychosis</u> | <u>one time or repeated psychedelic use</u> | <u>Bipolar history or family history?</u> | <u>Additional factors</u> | <u>Did the symptoms resolve?</u> |
| --- | --- | --- | --- | --- | --- | --- | --- | --- | --- |
| 1 | Cohen, 1966 | Unclear | LSD | none indicated | mania and possible psychosis | repeated | No | time line is unclear | Not stated |
| 2 | Reich & Hepps, 1972 | 8-10 days? | LSD | prior history of marijuana, LSD & amphetamine use | psychosis and mania | one time use for the current episode; prior use | No |  | 4 months |
| 3 | Hyde et al., 1978 | 5 - 10 days | psilocybin | none indicated | psychosis and possible mania | multiple times over course of a week plus large dose | No previous diagnosis or family history noted | very large dose | After 10 days |
| 4 | Davies, 1979 | hospitalized for 5 weeks | psilocybin | no; polysubstance use history | psychosis and possible mania | repeated | No previous diagnosis or family history noted | not clear that symptoms tied to mushroom use | 5 weeks, but continued depression |
| 5 | Dewhurst, 1980 | hospitalized for 10 weeks | psilocybin | none indicated during episode but frequent polysubstance use | both mania and psychosis | one time large dose; followed by separate doses | No previous diagnosis or family history noted | very large dose | Possibly at 10 weeks |
| 6 | Lake et al., 1981 | 6 weeks | LSD | alcohol | Mania | one time | No |  | After 3 weeks |
| 7 | Perera, Ferraro, & Pinto, 1995 | 6 weeks to 3 months | LSD | none indicated | both mania and psychosis | one time | family history of bipolar disorder & schizophrenia |  | Unclear, lost to follow up |
| 8 | Bickel et al., 2005 | 1 month, possibly longer | psilocybin | no; polysubstance abuse use history | psychosis and possible mania | one time | No previous diagnosis or family history noted | long history of heroin, opiate, and cannabis abuse; unclear from description if symptoms are definitely mania | At 6 month visit no symptoms |
| 9 | Paterson, Darby, & Sandhu, 2015 | 21 days | DMT | concurrent cannabis use | possibly both | repeated DMT use | family history of bipolar |  | psychosis resolved 2-3 weeks after admission; 6 month follow up visit showed no |
| 10 | Sami et al., 2015 | 6 months | DMT, LSD, other | cannabis, ketamine, DMT, LSD, Salvia Divinorum, synthetic cannabinoids, and cocaine | Both | unclear except for 'heavy user of psychedelics' | No | TBI & heavy polysubstance use | No |
| 11 | Szmulewicz, Valerio, & Smith, 2015 | 1 month+ | ayahuasca | none indicated | mania and psychosis | unclear | previous hypomanic-like episodes; father had bipolar I |  | asymptomatic at 1 month |
| 12 | Prajapati et al., 2016 | 12 days | LSD | cannabis; possible polysubstance use | psychosis; possible mania | repeated | No | Not clear that symptoms came from LSD use | symptoms improved at 6 days; no symptoms at 10 months |
| 13 | Brown et al., 2017 | 7 days | DMT | DMT and phenelzine | both mania and psychosis | multiple uses of DMT | Bipolar I |  | Unresolved, patient left AMA |
| 14 | dos Santos, Bouso, and Hallak, 2017 | 2-4 days | ayahuasca | history of MDMA & marijuana use | psychosis, possibly mania | two times, once per day in two sequential days | No |  | symptoms mostly resolved over months; 1 year follow up no symptoms |
| 15 | Barbic et al., 2020 | 20 days | DMT | none indicated; regular use of alcohol and cannabis | Psychosis and possible mania | one time | First cousin with bipolar disorder | Timeline is unclear | At 20 days |
| 16 | Hendin and Penn, 2021 | 1 week | psilocybin | cannabis, but not immediately proximal to mania | psychosis and mania | one time | father and paternal grandmother with bipolar |  | about 1 week |
| 17 | Palma-Alvarez et al., 2021 | >10d (exact length unspecified) | ayahuasca | none indicated | psychosis & possible mania | repeated | No previous diagnosis or family history noted | history of cocaine use disorder noted | Yes, following antipsychotic medication use (exact timeframe not mentioned) |

First, one of our primary goals was to find clear examples of individuals with a diagnosis of bipolar disorder (or previous history of manic/hypomanic symptoms) who had an adverse outcome after taking a psychedelic substance. Of the 17 cases that met the broader criteria, we found two such cases. In Case 13, a psychiatrist with bipolar disorder type I was hospitalized for mania after self-medicating his depression with 1g of vaporized DMT daily for 6 months and 60mg of phenelzine daily for three months (Brown et al., 2017). It is unclear if his manic episode was precipitated by factors such as the combination of DMT and an MAOI, time frame, or one of the distinct substances. In Case 11, a 30-year-old man developed symptoms of mania, delusions, and hallucinations two days after participating in a four-day ayahuasca retreat (Szmulewicz et al., 2015). Notably, this individual had symptoms of hypomania (but no depression) two weeks prior to the retreat. The case specifically noted that he was at the retreat to learn about the culture and services, and not as a specific treatment. The presence of hypomania before the retreat raises the possibility that issues related to developing mania (e.g., increasing impulsivity) may have led to his psychedelic use and that he may have developed mania even without using a psychedelic.

Another primary goal was to find cases where an individual without a prior history of bipolar disorder took a psychedelic substance (without taking other substances) that appeared to activate a manic episode. We found four cases where an individual with no known bipolar history (although two cases had a family history of bipolar disorder) took a single psychedelic substance once (without any other substance or a polysubstance use history), which resulted in sustained manic symptoms. Case 6 was a 31-year-old man who took LSD one time and two weeks later experienced dramatically decreased sleep along with an increased focus on religious themes and broke into a neighbor’s house. He was hospitalized and treated successfully with lithium (Lake et al., 1981). In Case 7, a 21-year-old woman was admitted to a psychiatric hospital with persistent manic and psychotic symptoms one week after taking LSD at a music festival (Perera et al., 1995). At admission she had decreased need for sleep, paranoia, mood swings, and catatonia. Her mother had a diagnosis of bipolar disorder, and her uncle a diagnosis of schizophrenia. Case 2 describes a 22-year-old man who took LSD, became paranoid, flew to his home in Israel, took all of his money out of the bank, then flew back and traveled throughout Europe, convinced that he was escaping a Nazi conspiracy (Reich and Hepps, 1972). This ultimately led him to attack and kill a stranger that he thought was a Nazi soldier. He was hospitalized for four months before his psychosis and manic symptoms remitted. Finally, Case 16 describes a 21-year-old woman who presented to outpatient psychiatry one month following hospitalization related to an episode of mania triggered by psilocybin use (Hendin & Penn, 2021). She reported that while her psilocybin experience was pleasant and “mystical”, 36 hours after dosing she experienced irritability, pressured speech, decreased need for sleep, paranoia, and delusions that led her family to contact the police. She was admitted to inpatient behavioral health and stabilized on lithium and aripiprazole, and later lamotrigine. On follow-up, she had one irritable mixed episode of mania that was not related to recent psychedelic use that resolved. While the overall number of these cases is small, they highlight that a manic episode can occur when individuals take a psychedelic substance, perhaps especially when one is predisposed to bipolar disorder.

Another important point is that 13 of the 17 cases found involved recreational psychedelic use, leaving three exceptions: Case 13 of the psychiatrist who was self-medicating with DMT (Brown et al., 2017), Case 11 of the 30-year-old man with a history of hypomania who participated in a four-day ayahuasca retreat (Szmulewicz et al., 2015), and Case 14 of a 40-year-old woman who participated in a two-day ayahuasca retreat (Lima and Tofoli, 2011). In this third case, the patient began to exhibit psychotic symptoms on the second day of the retreat and subsequently had two bouts of sleeplessness (including one 48-hour period), delusional beliefs, and excessive talking. The symptoms remitted after she was hospitalized and treated with haloperidol, and she reported no additional symptoms on follow-up one year later. Notably, these last two cases are the only two cases that we found where the individuals had clear manic episodes after taking a psychedelic substance as a treatment in a guided session. It is unclear whether this low number reflects the fact that those performing unregulated treatments screen out individuals with histories that may indicate susceptibility to mania or psychosis, if proper attention to set and setting decreases these outcomes, if these issues are just not well documented, or some combination of these factors. Unfortunately, the ayahuasca ceremony cases identified do not describe the specifics of the set and setting of the ritual retreat, which may be important to understanding the risk of activation of mania in this population.

Finally, it should be noted that in our screening of the literature, we identified a number of published cases that include clear serious adverse outcomes (e.g., severe medical accidents, death, and possible suicides) during the period of intoxication. However, we did not find any published cases where an individual had a diagnosis of bipolar spectrum disorder, took a psychedelic substance, and then died due to an accident or suicide.

## DISCUSSION

Overall, the present review focused on the available evidence of risk that psilocybin confers in bipolar disorder. We found several published case studies that implicate psychedelic use as a possible contributor to a manic episode. As described in Table 1, we found 17 cases that describe possible activation of a manic episode (or a psychotic episode that may have had manic elements) after psychedelic use. Of those cases, two (Case 11 and 13) appeared to be people who may have had a bipolar disorder diagnosis prior to taking the substance, and five had a family history of bipolar disorder. Other important themes that our case review indicates is that polysubstance use (5 of 17 cases) and the use of psychedelics multiple times over a short time period (9 of 17 cases) are both risk factors for adverse psychiatric outcomes. Together, this suggests that several factors related to psychedelic use may contribute to the risk of a manic episode in this population.

Though the cases are significant, the small number of published case studies linking psychedelic use to mania induction is not overwhelming. Furthermore, the number of cases in which ingestion of psilocybin led to a possible manic episode is surprisingly small (n=5) and only one case involved someone without a history of bipolar disorder developing mania after a clear single ingestion of a psychedelic involved psilocybin or hallucinogenic mushrooms (Case 16). Given that psychedelic use in general, and ingestion of psilocybin-containing mushrooms in particular, are common recreationally (Center for Behavioral Health Statistics and Quality, 2016) and in unregulated treatment settings, this suggests that the overall rate of inducing mania may be relatively low. Of course, it may be that the small number of published case studies is due to perception (or reality) that examples of mania or worsening of bipolar disorder after psychedelic use are so common that they do not warrant publication as case studies. However, this seems unlikely given the epidemiological research indicating that recreational use of psychedelic substances does not appear to be an independent risk factor for mania or psychosis (Johansen and Krebs, 2015). While the recreational use of psychedelics in the general population is certainly substantive, the number of individuals with bipolar disorder (or at risk for the disorder) taking a psychedelic is not known, and so the actual risk is not known.

Between this review of the published case study literature, the exclusion of individuals with bipolar disorder in modern trials, and the epidemiological data on hallucinogen use, perhaps the most striking finding is how little is known about the effects of psilocybin and similar psychedelics in this patient population. It is evident from this review that much more research is needed, including direct evaluations of psychedelic experience and outcomes among individuals with bipolar disorder, in both recreational and unregulated treatment settings. For example, researchers could directly solicit responses from individuals with bipolar disorder on their experiences with psychedelic substances, or from providers or guides in unregulated (or newly regulated) treatment settings. It is certainly possible that only a small percentage of individuals with bipolar disorder (or at risk for the disorder) would develop a manic episode or worsening of the disorder on exposure to a psychedelic substance.

While this review has focused on the potential risks for individuals with bipolar disorder, it may be that some patients could benefit from psychedelic treatments. In our search there was one case study (not included in the 17 cases) that implicated an accidental LSD overdose in the remission of bipolar disorder for over 20 years (Haden and Woods, 2020). A 15-year-old girl (monitored closely by mental health providers following diagnosis of bipolar disorder with psychotic features at age 12), accidentally took 10 times the recreational dosage of LSD and showed essentially no subsequent psychotic, hypomanic, or manic symptoms for 20 years after the event. The adolescent’s parent, psychiatrist, and therapist followed her closely over the subsequent years and attributed her mental health stability to the overdose. We did not find other similar cases in our review, and clearly this case does not provide the kind of evidence needed to evaluate psychedelics in the treatment of bipolar disorder. It is also possible that a psychedelic drug (such as psilocybin) may be especially challenging for people with bipolar disorder, given the feelings of euphoria that often accompany use. Specifically, there is some evidence that people with bipolar disorder can develop a fear of strong positive feelings due to association with a manic episode, leading some researchers to include positive mood exposures into psychosocial treatments (Johnson and Fulford, 2008).

There are several reasons that caution around psilocybin use is indicated in this population including: the strong serotonergic activation of psychedelic substances and the possibility of a TEAS, the potential for adverse events in a population known for impulsive behavior, and clinical case reports of negative events. Because all modern clinical psychedelic trials have excluded people with bipolar disorder, we lack critical information about the effects of psilocybin and similar compounds in this clinical population. However, people with bipolar disorder carry a significant burden of depression that may respond positively to the antidepressant effects of psilocybin when carefully administered in a controlled setting. Given our findings here that the evidence for the activation of mania is not overwhelming after psychedelic use, this possibility deserves further exploration.

To assess the relative risk of activating a manic episode in bipolar (or worsening the disorder), the gold standard approach would be a prospective study. We, and others (e.g., https://clinicaltrials.gov/ct2/show/NCT04433845), are currently developing such a study, structured to mitigate potential risks in this population. First, we plan to optimize both the ‘set’ and the ‘setting’ in our clinical psilocybin trials. These aspects were missing in nearly every case report we reviewed. Second, we will screen patients for relevant risk factors, such as substance use and a history of psychosis. Third, we will use a conservative dose-escalation protocol, administering a low dose of psilocybin followed weeks later with a high dose in order to monitor participants’ responses over time. Fourth, we will initially only include individuals without a history of mania (i.e., individuals with bipolar 2) and individuals who are outside of the age range where a manic episode would likely first occur (i.e., >30 years old). Finally, we will require participants to have both community and psychological support in place to manage any lingering effects of the intervention after the study is completed. Using all of these precautions will likely significantly mitigate the risk of adverse events such as those reviewed here. Of course, given that it is possible that only a small percentage of people with bipolar disorder could develop a manic episode due to psychedelic use, ultimately very large prospective studies will need to be developed. Further, our treatment approach is initially designed for individuals with bipolar 2 disorder, which may be quite distinct from individuals with a history of mania. Subsequent studies may focus on individuals with bipolar 1, although at this stage, the available evidence suggests that this would be most likely focused on those with bipolar 1 with current depression.

There are several limitations to the case reviews that should be mentioned. First, it must be noted that we were limited to only those case studies that clinicians and researchers chose to submit for publication. This of course differs from published research that may include null results. Also, as mentioned above, it is certainly possible that there is a perception that experiences of mania brought on by psychedelic use is commonplace and thus not something that treatment providers would think to publish. Indeed, a review of published cases like this can only tell us what can happen, not how often these events occur. There were also several challenges in reviewing the case study literature and we were obviously limited to only the data provided in each individual case. Case details were often sparse, the motivation for the case review was not always on the outcome of the patient or the patient’s background (e.g., some were focused on urine and blood work assessments), timelines were frequently unclear, and most importantly there was often minimal follow-up. Because many of the published cases involved recreational psychedelic use, details leading up to the adverse events were often vague or hard to follow. Furthermore, the dosages (or confirmation of the substance) were frequently unavailable or reliant on patient reports. Additionally, approximately half of the cases involved either concurrent polysubstance use or recent polysubstance use; the role that drug interactions may have played is difficult to determine. Another limitation is that we focused specifically on persistent adverse events. There were a number of cases that did not involve a clear persistent event because the outcome resulted in the death of the patient (either from accident or suicide). It is possible that some of these cases involved individuals with undiagnosed bipolar spectrum disorders, and the psychedelic substance led to impulsive actions resulting in the patient’s death. However, this is purely speculative.

## Conclusion

This case review revealed a low number of cases of persistent manic symptoms that appeared to be brought on by use of a psychedelic substance, which may suggest that the rate of psychedelic-induced mania is low. However, not knowing the actual number of people who have bipolar disorder who have taken a psychedelic substance tempers this conclusion. Nonetheless, given the existing clinical and basic research, epidemiological studies, and published case histories, and importantly the lack of treatment options for depressive symptoms in bipolar disorder, we believe that a cautious approach to conducting trials in bipolar depression with psilocybin therapy, to examine the effects and safety is warranted. In such a prospective trial, it is crucial that attention be given to a carefully regulated environment where researchers can screen participants, emphasize set and setting, and follow patients for an extended period.

## Data Availability

Not applicable

## Notes

Sources of direct funding, support, or sponsorship Dr. Woolley is funded in part via VA CSR&D 1IK4CX002090-01. Grant title: Quantifying and Treating Social Deficits in Veterans with Mental Illness: a Five Year Plan.

Potential conflicts of interest Dr. Woolley is a paid consultant for Psilo Scientific Ltd. and Silo Pharma.

### Competing Interest Statement

Dr. Woolley is a paid consultant for Psilo Scientific Ltd. and Silo Pharma.

### Funding Statement

Dr. Woolley is funded in part via VA CSR&D 1IK4CX002090-01. Grant title: Quantifying and Treating Social Deficits in Veterans with Mental Illness: a Five Year Plan.

### Author Declarations

This is a review of the literature and no IRB approval was required.

### Summary of Updates

Change in the paper content including a search of published cases through 6/30/2021.

